# Pre-existing T cell memory as a risk factor for severe COVID-19 in the elderly

**DOI:** 10.1101/2020.09.15.20188896

**Authors:** Petra Bacher, Elisa Rosati, Daniela Esser, Gabriela Rios Martini, Carina Saggau, Esther Schiminsky, Justina Dargvainiene, Ina Schöder, Imke Wieters, Yascha Khodamoradi, Fabian Eberhardt, Holger Neb, Michael Sonntagbauer, Maria J.G.T. Vehreschild, Claudio Conrad, Florian Tran, Philip Rosenstiel, Robert Markewitz, Klaus-Peter Wandinger, Jan Rybniker, Matthias Kochanek, Frank Leypoldt, Oliver A. Cornely, Philipp Koehler, Andre Franke, Alexander Scheffold

**Affiliations:** Institute of Immunology, Christian-Albrechts-University of Kiel & UKSH Schleswig-Holstein, Kiel, Germany; Institute of Clinical Molecular Biology, Christian-Albrechts-University of Kiel, Kiel, Germany; Neuroimmunology, Institute of Clinical Chemistry, University Hospital Schleswig-Holstein, Kiel, Germany; Department of Internal Medicine, Infectious Diseases, University Hospital Frankfurt & Goethe University Frankfurt, Frankfurt am Main, Germany; Department of Anesthesiology, Intensive Care Medicine and Pain Therapy, University Hospital Frankfurt, Frankfurt am Main, Germany; Department of Internal Medicine, Hospital of Preetz, Preetz, Germany; Department of Internal Medicine I, UKSH Kiel, Germany; Institute of Clinical Chemistry, University Hospital Schleswig-Holstein, Lübeck, Germany; University of Cologne, Medical Faculty and University Hospital Cologne, Department I of Internal Medicine, 50937 Cologne, Germany; Center for Molecular Medicine Cologne (CMMC), University of Cologne, 50931, Cologne, Germany; University of Cologne, Medical Faculty and University Hospital Cologne, German Center for Infection Research (DZIF), Partner Site Bonn-Cologne, 50937 Cologne, Germany; Clinical Trials Centre Cologne, ZKS Köln, 50935 Cologne, Germany; University of Cologne, Cologne Excellence Cluster on Cellular Stress Responses in Aging-Associated Diseases (CECAD), 50937 Cologne, Germany; Lead contact

## Abstract

Coronavirus disease 2019 (COVID-19) displays high clinical variability but the parameters that determine disease severity are still unclear. Pre-existing T cell memory has been hypothesized as a protective mechanism but conclusive evidence is lacking. Here we demonstrate that all unexposed individuals harbor SARS-CoV-2-specific memory T cells with marginal cross-reactivity to common cold corona and other unrelated viruses. They display low functional avidity and broad protein target specificities and their frequencies correlate with the overall size of the CD4+ memory compartment reflecting the “immunological age” of an individual. COVID-19 patients have strongly increased SARS-CoV-2-specific inflammatory T cell responses that are correlated with severity. Strikingly however, patients with severe COVID-19 displayed lower TCR functional avidity and less clonal expansion. Our data suggest that a low avidity pre-existing T cell memory negatively impacts on the T cell response quality against neoantigens such as SARS-CoV-2, which may predispose to develop inappropriate immune reactions especially in the elderly. We propose the immunological age as an independent risk factor to develop severe COVID-19.

**Key points:**

- Pre-existing SARS-CoV-2-reactive memory T cells are present in all humans, but have low functional avidity and broad target specificities
- Pre-existing memory T cells show only marginal cross-reactivity to common cold corona viruses
- Frequencies of pre-existing memory T cells increase with the size of the CD4+ memory compartment reflecting the “immunological age” of the individual
- Low-avidity and polyclonal, but strongly enhanced SARS-CoV-2 specific T cell responses develop in severe COVID-19, suggesting their origin from pre-existing memory
- The immunological age may represent a risk factor to develop severe COVID-19

## Introduction

COVID-19 displays remarkable disparity of clinical symptoms, ranging from asymptomatic or mild disease frequently observed in children and younger adults to severe clinical symptoms associated with high mortality mainly in elderly and high-risk patients. Differences in the immune response may contribute to this diverse pathology. Severe disease is characterized by hyperinflammation, suggesting that exaggerated immune reactions are part of COVID-19 pathogenesis. However, it is currently not clear which type of adaptive immunity to SARS-CoV-2 is protective or detrimental. Thus, there is an enormous interest to decipher the anti-SARS-CoV-2 response, both to define parameters of immune protection *versus* pathology, as well as for the design of effective vaccination strategies.

SARS-CoV-2-specific CD4+ T cells are prime candidates to be involved in this process. They are central organizers of anti-viral immune responses while uncontrolled T cell responses may cause pathology. Severe lymphopenia accompanies severe disease and T cell reappearance correlates with patient recovery (Huang et al., 2020; Tan et al., 2020; Wang et al., 2020; Yang et al., 2020). Markers of T cell activation were found to be increased on total (Diao et al., 2020; Sekine et al., 2020; Wilk et al., 2020; Zheng et al., 2020), as well as on SARS-CoV-2-specific T cells (Braun et al., 2020; Sekine et al., 2020). Overall COVID-19 patients seem to develop robust Th 1 -like SARS-CoV-2-specific CD4+ T cell responses focused on spike, membrane and nucleocapsid (Ncap) proteins (Grifoni et al., 2020). Increased frequencies of SARS-CoV-2-specific T cells have been correlated with more severe disease (Anft et al., 2020; Peng et al., 2020) supporting the idea that exaggerated CD4+ T cell responses may contribute to the hyperinflammation. However, the factors which determine the magnitude as well as the quality of the CD4+ T cell response and how this relates to predisposition and/ or manifestation of severe disease remains unknown. In particular, the effect of aging is discussed, since the risk to develop severe COVID-19 dramatically increases in the elderly.

Several studies have observed that a certain fraction of un-exposed donors have pre-existing SARS-CoV-2-reactive T cells (Braun et al., 2020; Grifoni et al., 2020; Le Bert et al., 2020; Mateus et al., 2020; Meckiff et al., 2020; Sekine et al., 2020; Weiskopf et al., 2020) which contained at least some T cells cross-reactive against selected peptides with homology to related common cold corona virus strains (CCCoV) (Braun et al., 2020; Mateus et al., 2020). From this it was hypothesized that encounter with CCCoV may provide protective cross-reactive memory especially in younger patients, where infections with CCCoV are especially prevalent.

However, data on the prevalence of CD4+ T cell responses against CCCoV in humans are lacking. Furthermore, pre-existing immunity has also been described for several other pathogens and neoantigens (Bacher et al., 2013; Campion et al., 2014; Kwok et al., 2012; Su et al., 2013) with variable consequences, from protective to harmful (Bacher et al., 2019; Greiling et al., 2018; Koutsakos et al., 2019; Sridhar et al., 2013; Welsh et al., 2010; Woodland and Blackman, 2006). Thus its impact may depend on the T cells functional characteristics, the specific antigen- or pathogen-context (Sette and Crotty, 2020), and age (Woodland and Blackman, 2006). Such functional characteristics of SARS-CoV-2-specific T cells in severe *versus* mild COVID-19 and unexposed individuals are still poorly described. Specifically, the prevalence of the putative cross-reactive T cells within unexposed donors and COVID-19 patients and in different age groups, their phenotypic and functional characteristics, as well as the inducing antigen(s) are unknown.

Here we show that pre-existing memory T cells are present in all unexposed donors and increased in the elderly, but not primarily driven by CCCoVs. Pre-existing SARS-CoV-2-specific memory T cells possess only low TCR avidity, suggesting impaired functionailty. This functional impairment is closely mirrored in T cells from severe COVID-19 patients in contrast to mild disease, suggesting that they may originate from pre-existing memory T cells. Thus we suggest the immunological age as a potential risk factor for severe COVID-19.

## Results

### Strongly increased frequencies of human SARS-CoV-2-reactive CD4+ T cells against the spike, membrane and Ncap proteins in COVID-19 patients

To characterize the human T cell response against SARS-CoV-2, we analyzed T cells reactive against a panel of 12 different SARS-CoV-2 proteins. SARS-CoV-2-reactive CD4+ T cells were detected based on the up-regulation of CD154+ (CD40L) following 7h ex vivo stimulation of PBMCs with overlapping peptide pools of the different proteins and subsequent magnetic enrichment (Antigen-reactive T cell enrichment, ARTE) (Bacher et al., 2016; Bacher et al., 2019) (Figure S1A). SARS-CoV-2 exposure *versus* non-exposure of blood donors was verified by SARS-CoV-2 PCR and/ or serology testing (Table S1).

The response of COVID-19 patients was mainly directed against three proteins, spike, membrane and nucleocapsid (Ncap), as previously suggested (Grifoni et al., 2020), as well as to lower extent and with more variability between donors against AP3a, ORF9b, NS6, NS7a and NS8 (Figure 1A, B). We observed no differences in the reactivity against the N-terminal or C-terminal part of the spike protein in COVID-19 patients. The frequencies of reactive cells against single or pooled spike, membrane and Ncap, were strongly increased in patients *versus* unexposed individuals (Figure 1C), whereas no differences were detected against a pool of Influenza A H1N1 proteins (containing HA, MP1, MP2, NP and NA), as a control antigen. In contrast to previous reports suggesting pre-existing memory only in a subset of unexposed individuals, the sensitive detection by ARTE identified SARS-CoV-2 reactive T cells in all unexposed donors albeit at low and variable frequencies ranging from 1 in 10^-5^-10^-3^ (Figure 1A-C). However, while typically >80-90% of SARS-CoV-2 reactive T cells in COVID-19 patients were directed against spike, membrane and Ncap, the response in unexposed donors was much more variable and directed against multiple proteins (Figure 1D) (Grifoni et al., 2020; Le Bert et al., 2020). The specificity of the SARS-CoV-2-reactive cells in unexposed as well as exposed donors was confirmed by high reactivity of sorted and expanded CD154+ T cells towards SARS-CoV-2, but not control antigens (Figure S1B, C).

**Figure 1.**
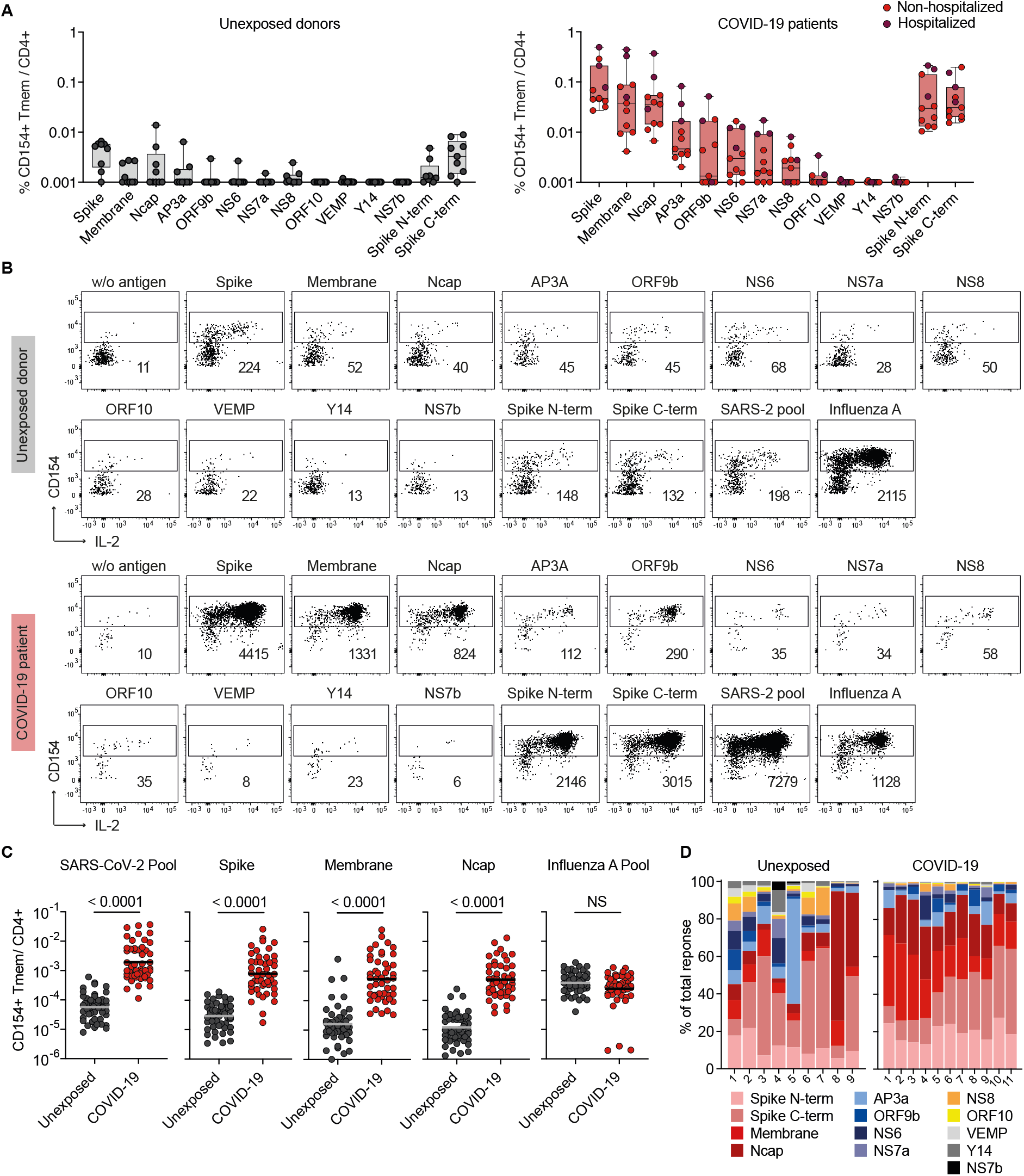
Identification of immunogenic SARS-CoV-2 proteins. (A) Frequencies of reactive CD154+CD45RA-memory CD4+ T cells (Tmem) against individual SARS-CoV-2 proteins in unexposed donors (n=9) and COVID-19 patients (n=11; non-hospitalized n=8; hospitalized n=3). (B) Representative dot plot examples for *ex vivo* detection of SARS-CoV-2-reactive CD4+ T cells by ARTE. Absolute cell counts after magnetic CD154+ enrichment from 1×10e7 PBMCs are indicated. (C) Frequencies of SARS-CoV-2-reactive Tmem against individual or pooled spike, membrane, Ncap proteins or a pool of Influenza A proteins (containing HA, MP1, MP2, NP and NA). Unexposed donors (n=50), COVID-19 patients (n=49). (D) Proportion of SARS-CoV-2 proteins recognized by CD4+ T cells in unexposed donors (n=9) and COVID-19 patients (n=11). Each symbol in (A, C) represents one donor. (A) Box-and-whisker plots display quartiles and range. (C) Horizontal lines indicate geometric mean. Statistical differences: (C) Two-tailed Mann-Whitney test.

### SARS-CoV-2 reactive T cells of COVID-19 patients show an activated Th1/Tfh-like signature

SARS-CoV-2 reactive cells from COVID-19 patients *versus* unexposed individuals displayed increased expression of the acute and chronic activation markers Ki-67 and CD38 (Figure 2A), as reported by others (Braun et al., 2020; Sekine et al., 2020). The expression of both markers declined with time after infection, but not the frequencies of reactive T cells (Figure 2B, C). We also detected slightly increased relative and strongly increased absolute production of inflammatory cytokines in COVID-19 patients, such as IL-2, IFN-γ and IL-21 compared to unexposed donors, as well as a slightly higher production of IL-10 (Figure 2D, E). While inflammatory cytokines increased with time after infection, IL-10 was mainly produced during active disease (Figure S2A), suggesting a counter-regulatory mechanism during acute infection. In addition, SARS-CoV-2 reactive T cells expressed stably high levels of PD-1 (CD279) (Figure 2D, E and S2A). Compared to other anti-viral reponses, production of TNF-α, IFN-γ and IL10 was rather reduced in convalescent COVID-19 patients, while IL-21 and PD-1 were highly increased (Figure 2F, Figure S2B). We observed no differences in the cytokine response or phenotype between the individual SARS-CoV-2 proteins (Figure 2F, Figure S2B).

**Figure 2.**
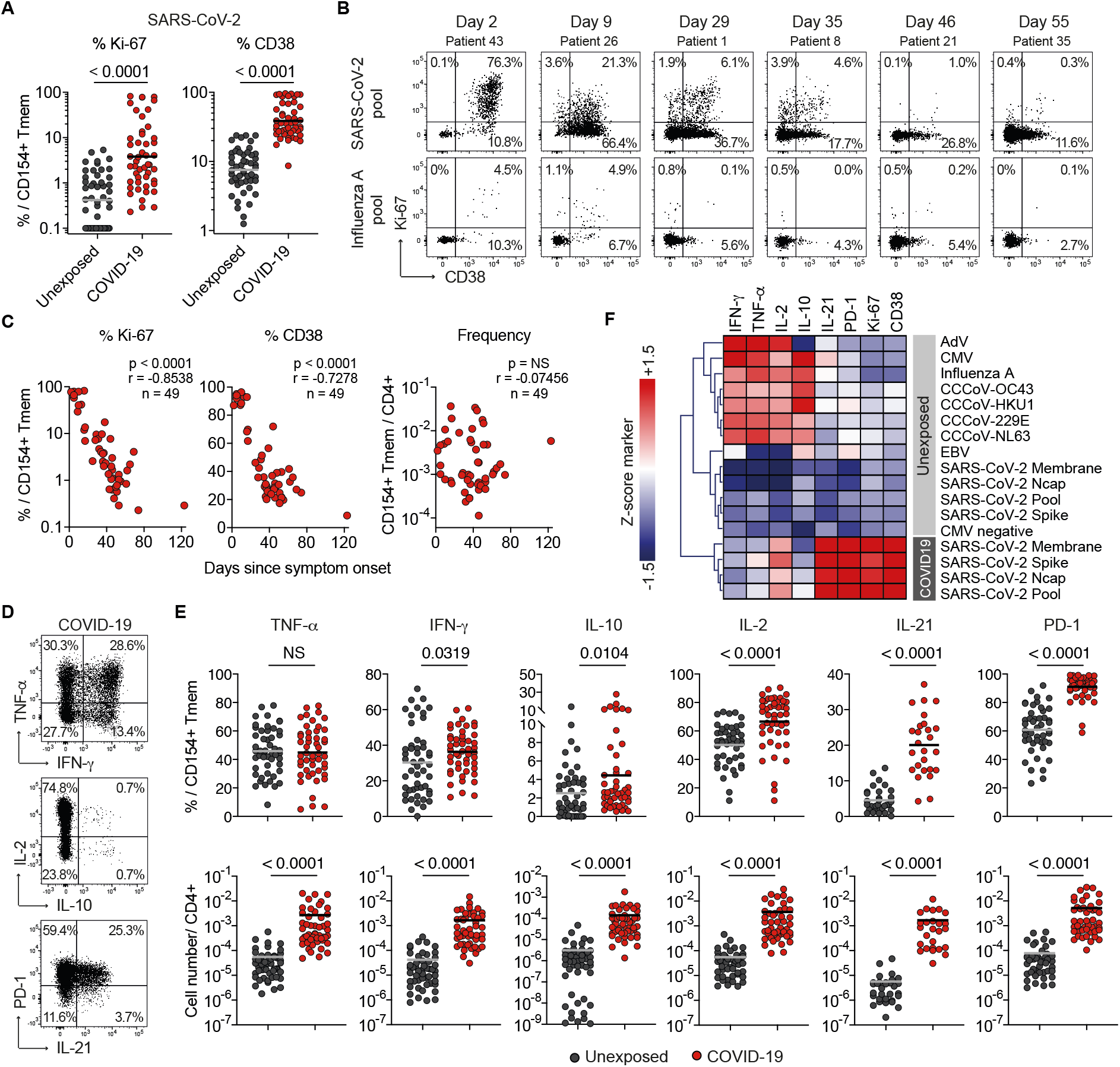
Inflammatory SARS-CoV-2-specific CD4+ T cell responses in COVID-19 patients. (A) *Ex vivo* Ki-67 and CD38 expression of SARS-CoV-2 pool-reactive CD154+ Tmem. Unexposed donors (n=50), COVID-19 patients (n=49). (B) *Ex vivo* Ki-67 and CD38 staining of SARS-CoV-2 pool- or Influenza A-reactive CD154+ Tmem from COVID-19 patients at different time points after disease onset. Percentage of Ki-67+ and/ or CD38+ cells within CD154+ Tmem are indicated. (C) Spearman correlation of Ki-67 and CD38 expression or frequencies of SARS-CoV-2 pool-reactive CD154+ Tmem and days since disease onset in COVID-19 patients (n=49). (D) *Ex vivo* cytokine and phenotype staining of SARS-CoV-2 pool-reactive CD154+ Tmem from a COVID-19 patient. Percentage of marker positive cells within CD154+ Tmem are indicated. (E) *Ex vivo* cytokine production and phenotype of SARS-CoV-2 pool-reactive cells. Upper row: within CD154+ Tmem and lower row: within total CD4+ T cells. Unexposed donors (n=50; IL-21 n=31), COVID-19 patients (n=49; IL-21 n=26). (F) Heatmap depicting *ex vivo* cytokine production of virus-reactive memory T cells (n=26-50). Cytokine production within CD154+ Tmem was measured by flow cytometry and mean values were Z score normalized for each cytokine. Each symbol in (A, C, E) represents one donor, horizontal lines indicate (A) geometric mean, (E) mean. Statistical differences: (A, E) Two-tailed Mann-Whitney test.

### Single-cell RNA sequencing identifies similar T cell clusters in COVID-19 and unexposed donors

To obtain a deeper insight into the cellular composition of SARS-CoV-2-specific T cells and their molecular patterns we next performed single-cell RNA sequencing of *ex vivo* FACS-purified SARS-CoV-2 reactive memory T cells. After quality filtering (see Methods) we analyzed in total 104,417 single cells from 6 unexposed and 14 COVID-19 patients.

UMAP cluster analysis revealed five clusters with a distinct transcriptional profile (Figure 3A). These were assigned as T follicular-helper-like (Tfh-like, key marker genes *IL21, POU2AF1*), transitional memory (*CD28, IL7R*), central memory (*CCR7, SELL*), cytotoxic (*IFNG, CSF2, PRF1, GNLY)*, type-I interferon response (*MX1, OAS1*) and cycling T cells (*MKI67, CDK1*) (Figure 3B). Similar clusters have recently been described in anti-viral T cells (Meckiff et al., 2020). However, especially Tfh-like, transitional and central memory T cells were related and important genes like *IFNG, CSF2, IL21, IL2* and *PDCD1* were expressed by many cells in all clusters although at different level (Figures 3B, Figure S3A). In addition, we observed three robust clusters, cytotoxic /Th1, type-I interferon, and cycling, which are indicative of cellular activation and an anti-viral type-I interferon response. These results confirm our cytometric analysis pointing to a highly activated Th1 and Tfh-like phenotype of SARS-CoV-2 specific T cells in COVID-19. However, similar clusters were also identified in SARS-CoV-2 reactive memory T cells from unexposed individuals and we were not able to clearly separate unexposed donors from COVID-19 patients or between patients with different disease severity only based on qualitative differences of the reactive T cells (Figure 3C, Figure S3B). There was a tendency that clusters indicative of acute activation, such as cycling and type-I interferon were relatively enriched in COVID-19 and Tfh cells were more abundant in mild COVID19 (Figure 3C).

**Figure 3.**
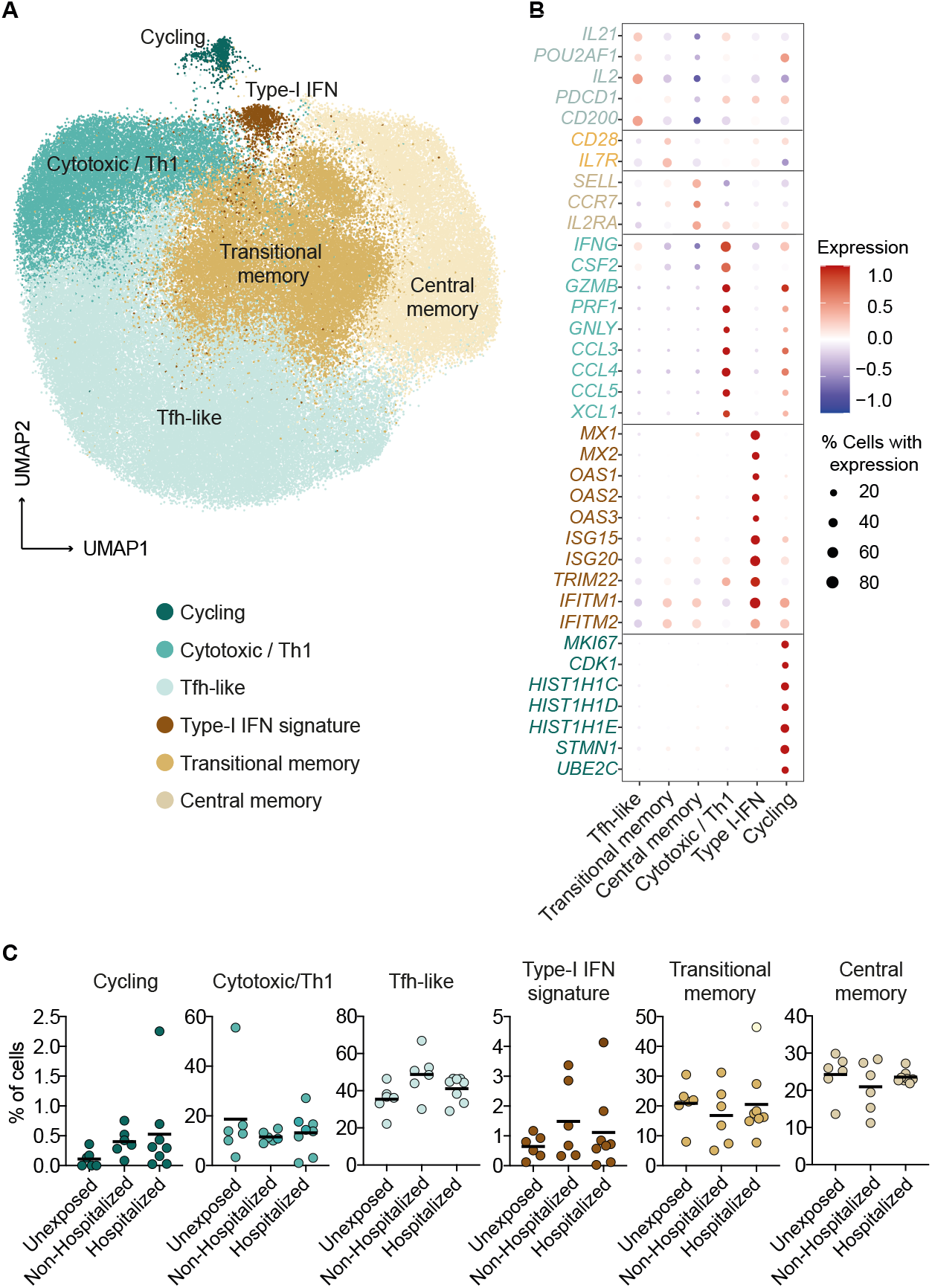
Single cell RNA sequencing of SARS-CoV-2-reactive CD4+ T cells. (A) Single cell gene expression of FACS purified *ex vivo* isolated CD154+ memory T cells following stimulation with pooled SARS-CoV-2 spike, membrane and Ncap proteins from unexposed donors (n=6) and COVID-19 patients (n=14). UMAP visualization of the subset composition of SARS-CoV-2 reactive CD4+ T cells colored by functional gene expression clusters. (B) Dot plot visualization showing the expression of selected marker genes in each SARS-CoV-2 T cell cluster. Colors represent the Z-score normalized expression levels and size indicates the proportion of cells expressing the respective genes. (C) Proportion of cells falling within each cluster for the individual donors (unexposed donors n=6; non-hospitalized COVID-19 patients n=6; hospitalized COVID-19 patients (n=8). Each symbol in (C) represents one donor, horizontal lines indicate mean.

Taken together, the cytometric and single cell sequencing data confirm that COVID-19 patients generate a strong pro-inflammatory Th1/cytotoxic-like and Tfh-like response against SARS-CoV-2 spike, membrane, and Ncap proteins. Interestingly though, the differences between the patients groups and healthy controls were mainly quantitative, rather than qualitative. This suggests that these cell types are not unique to COVID-19 but may represent a common cellular phenotype of anti-viral T cells, which are already present in pre-existing SARS-CoV-2-reactive memory T cells from healthy unexposed donors.

### Low avidity SARS-CoV-2-reactive memory T cells increase with age in unexposed donors

Recent studies have demonstrated pre-existing T cell immunity against SARS-CoV-2 presumably against common cold viruses in 20-50% of unexposed donors (Braun et al., 2020; Grifoni et al., 2020; Le Bert et al., 2020; Mateus et al., 2020; Meckiff et al., 2020; Sekine et al., 2020; Weiskopf et al., 2020). However, the antigen specificity as well as the clinical relevance of this cross-reactivity remains unknown (Sette and Crotty, 2020) although some cross-reactivity against homologous CCCoV epitopes has been found (Braun et al., 2020; Mateus et al., 2020). As shown in Figure 1, by sensitive enrichment of reactive T cells we detected low frequencies of cross-reactive T cells against different SARS-CoV-2 proteins in 100% of unexposed donors. To further characterize these pre-existing SARS-CoV-2-reactive T cells, we determined the proportion of memory *versus* naïve cells. Remarkably, a substantial fraction of SARS-CoV-2-reactive cells from unexposed donors but not COVID-19 patients displayed a naïve phenotype, as evidenced by high expression of CD45RA and CCR7 and lack of effector cytokine expression (Figure 4A, B; Figure 2F). The proportion of memory cells was highly variable between different donors (range 25-95%) (Figure 4B). It has further been speculated that this pre-existing immunity may improve protection especially in young patients and children due to frequent infections with common cold corona viruses (Braun et al., 2020). However, we detected no correlation of pre-existing T cell frequency with donor age (Figure 4C). Rather and in sharp contrast to the “pre-immune” hypothesis, the frequency (Figure 4D) and the proportion (Figure 4E) of SARS-CoV-2 cross-reactive memory cells of unexposed individuals positively correlated with the proportion of memory cells within the total CD4+ population that is associated to the immunological age. A similar pattern was observed for CMV-reactive T cells from CMV sero-negative *versus* sero-positive donors (Figure 4E), as well as T cells reactive against the neoantigen keyhole limpet hemocyanin (KLH) (Figure S4). These data argue against induction of the pre-existing SARS-CoV-2 memory cells by a specific cross-reactive antigen, but rather for arbitrary stochastic selection from a large memory repertoire in adult humans. In support of this, SARS-CoV-2-reactive memory T cells expanded from unexposed individuals displayed a 1-2 log lower functional avidity compared to COVID-19 patients, which was in the same range as CMV-reactive T cells (Figure 4F, G). Taken together, pre-existing SARS-CoV-2 cross-reactive memory T cells in unexposed donors are common in humans and increase with the immunological age but do not display features of a protective cross-reactive T cell population.

**Figure 4.**
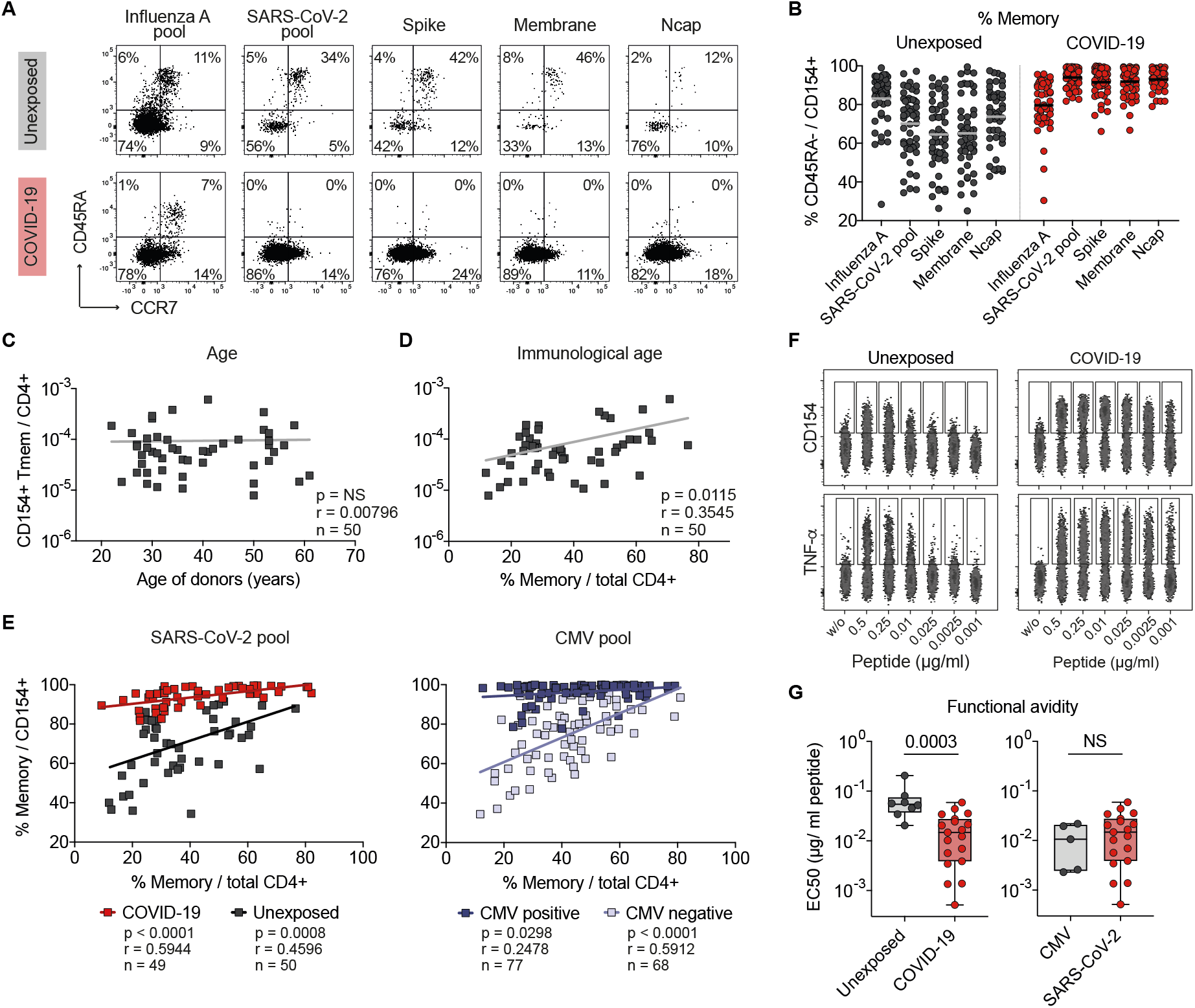
SARS-CoV-2 reactive CD4+ T cells in healthy donors. (A) CD45RA and CCR7 staining of SARS-CoV-2- or Influenza A-reactive CD154+ cells in unexposed donors or COVID-19 patients. Percentage of marker positive cells within CD154+ is indicated. (B) Proportion of memory cells within SARS-CoV-2 reactive cells in unexposed donors (n=50) or COVID-19 patients (n=49). (C, D) Spearman correlation between the frequencies of SARS-CoV-2 pool-reactive T cells in unexposed donors and (C) the age of donors or (D) the proportion of memory cells within the total CD4+ population, corresponding to the immunological age. (E) Pearson correlation between the proportion of memory cells within the antigen-specific T cells (y-axis) and the proportion of memory cells within the total CD4+ population (x-axis; immunological age) is shown for exposed and unexposed donors for SARS-CoV-2 and CMV. (F, G) SARS-CoV-2 pool-reactive CD154+ Tmem from unexposed donors and COVID-19 patients were FACS purified, expanded and re-stimulated with decreasing antigen concentration in the presence of autologous antigen-presenting cells. (F) CD154 or TNF-α expression for the indicated concentration per peptide. (G) EC50 values were calculated from dose-response curves. Left: SARS-CoV-2 reactive cells from unexposed donors n=8, COVID-19 patients n=19; right: CMV-reactive cells n=5 or SARS-CoV-2 reactive from COVID-19 patients (n=19). Each symbol in (B, C, D, E, G) represents one donor, horizontal lines indicate (B) mean. (G) Box-and-whisker plots display quartiles and range. Statistical differences: (G) Two-tailed Mann-Whitney test.

### Robust memory T cell response to common cold corona viruses (CCCoVs)

Our data do not exclude the possibility that in some donors protective pre-existing immunity may exist, for example due to infections with related common cold corona viruses (CCCoVs). Since data on the prevalence of CCCoV-specific T cell memory are lacking, we next analysed the response against spike proteins from the CCCoV strains 229E, OC43, HKU1 and NL63. Strikingly, robust memory T cell responses were readily detected in all donors with frequencies ranging between 1 in 10^3^-10^4^ (Figure 5A, B), which is in a similar range like against Influenza A (Figure 1C), but up to 10-fold higher than SARS-CoV-2 spike-reactive T cells (Figure 5B). CCCoV responses displayed a memory phenotype (Figure 5C, D), independent of immunological age (Figure S5A), and high functional avidity (Figure 5E) in accordance with an *in vivo* induction upon viral infection. Of note, expanded CCCoV-specific T cells from healthy donors showed only marginal cross-reactivity against SARS-CoV-2 spike protein and *vice versa* (Figure 5F, G). Furthermore, while the frequencies of reactive memory T cells between the different CCCoVs showed strong linear correlation as an indicator of cross-recognition, there was only a weak correlation between SARS-CoV-2 memory T cells and individual CCCoV strains (Figure 5H), which was in fact similar to other non-related common viral antigens (Figure S5B). To further analyse the potential relevance of pre-existing immunity to the anti-SARS-CoV-2 immune reponse, we re-stimulated expanded SARS-CoV-2-specific T cell lines from COVID-19 patients and unexposed donors (Figure 5I, J). Only minimal and highly variable cross-reactivity against CCCoV strains, but also against CMV or Influenza A were detected in COVID-19 patients, as well as in expanded cells from unexposed individuals, adding up to maximally 5% of the total response in individual donors (Figure 5I).

**Figure 5.**
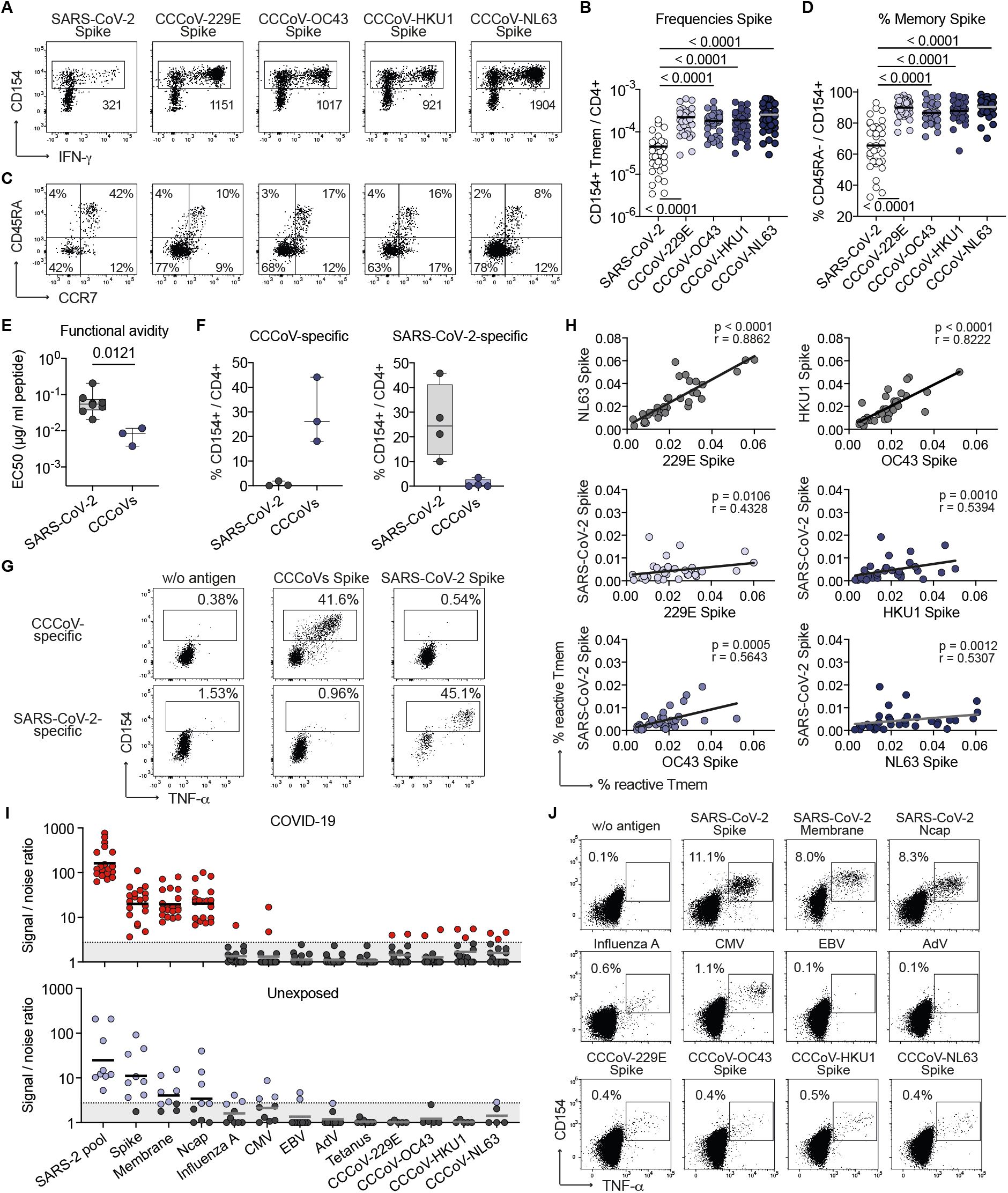
Human CD4+ T cell response against common cold viruses (CCCoVs) (A) *Ex vivo* detection of reactive CD4+ T cells against CCCoV spike proteins by ARTE. Absolute cell counts after magnetic CD154+ enrichment from 1×10e7 PBMCs are indicated. (B) Summary of CCCoV-reactive Tmem frequencies in healthy donors (n=34). (C) CD45RA and CCR7 staining of CCCoV-reactive CD154+ cells in healthy donors. Percentage of marker positive cells within CD154+ is indicated. (D) Proportion of memory cells within CCCoV-reactive cells in healthy donors (n=34). (E-G) CD154+ Tmem reactive against a pool of the 229E, OC43, HKU1 and nL63 spike proteins or reactive against the SARS-CoV-2 spike, membrane and Ncap proteins were FACS purified, expanded and re-stimulated. (E) Cells were re-stimulated with decreasing antigen concentration. EC50 values were calculated from dose-response curves. (F) Reactivity of the expanded cell lines against CCCoV spike pool or SARS-CoV-2 spike protein, respectively (n=3-4). (G) Representative dot plots for re-stimulation. Percentage of CD154+TNFa+ cells within CD4+ is indicated. (H) Spearman correlation between CD154+ Tmem frequencies reactive against different CCCoVs or CCCoVs and SARS-CoV-2 spike (n=34). (I, J) Expanded SARS-CoV-2 pool-reactive T cells from COVID-19 patients (n=19) or unexposed individuals (n=9) were re-stimulated with different antigens in presence of autologous antigen-presenting cells. (I) Signal:noise ratio of stimulated *versus* non-stimulated control. A detection limit (dashed line), was defined as signal:noise ratio ≥3. (J) Dot plot examples for re-stimulation of a COVID-19 patient. Cells were gated on CD4+ T cells and percentages of CD154+TNFα+ cells are indicated. Each symbol in (B, D, E, F, H, I) represents one donor, horizontal lines indicate (A, B) mean, (I) geometric mean. (E-F) Box-and-whisker plots display quartiles and range. Statistical differences: (B, D) Friedman test with Dunn’s post hoc test, (E) Two-tailed Mann-Whitney test.

Taken together these data clearly argue against a strong protective effect of pre-existing immunity in general and specifically against a major protective contribution of CCCoVs to the T cell response against SARS-CoV-2 in unexposed donors, as well as in COVID-19 patients.

### Increased, but unfocussed and low affinity CD4+ T cell response against SARS-CoV-2 in severe disease

Although we essentially excluded a general protective effect of pre-existing immunity we demonstrated that cross-reactive memory T cells against SARS-CoV-2 antigens are common in humans, increase with the immunological age and display rather low functional avidity. So far, the consequences of this stochastic pre-existing memory are unclear. Since elderly suffer more frequent from severe disease, we next compared the response of patients with mild *versus* severe disease. Classification was based on WHO criteria, whereby WHO groups 3-5 (moderate) and 6-7 (severe) were combined to increase statistical power (see Table S1). Interestingly, frequencies of reactive T cells against the single and pooled SARS-CoV-2 proteins, but not against Influenza A antigens positively correlated with disease severity (Figure 6A). This was not due to an age bias in severe disease as shown for a selected group of donors in the age range of 50-65 years (Figure 6B, C). Instead, we observed a clearly increased immunological age of hospitalized *versus* non-hospitalized patients within the same age group (Figure 6D).

**Figure 6.**
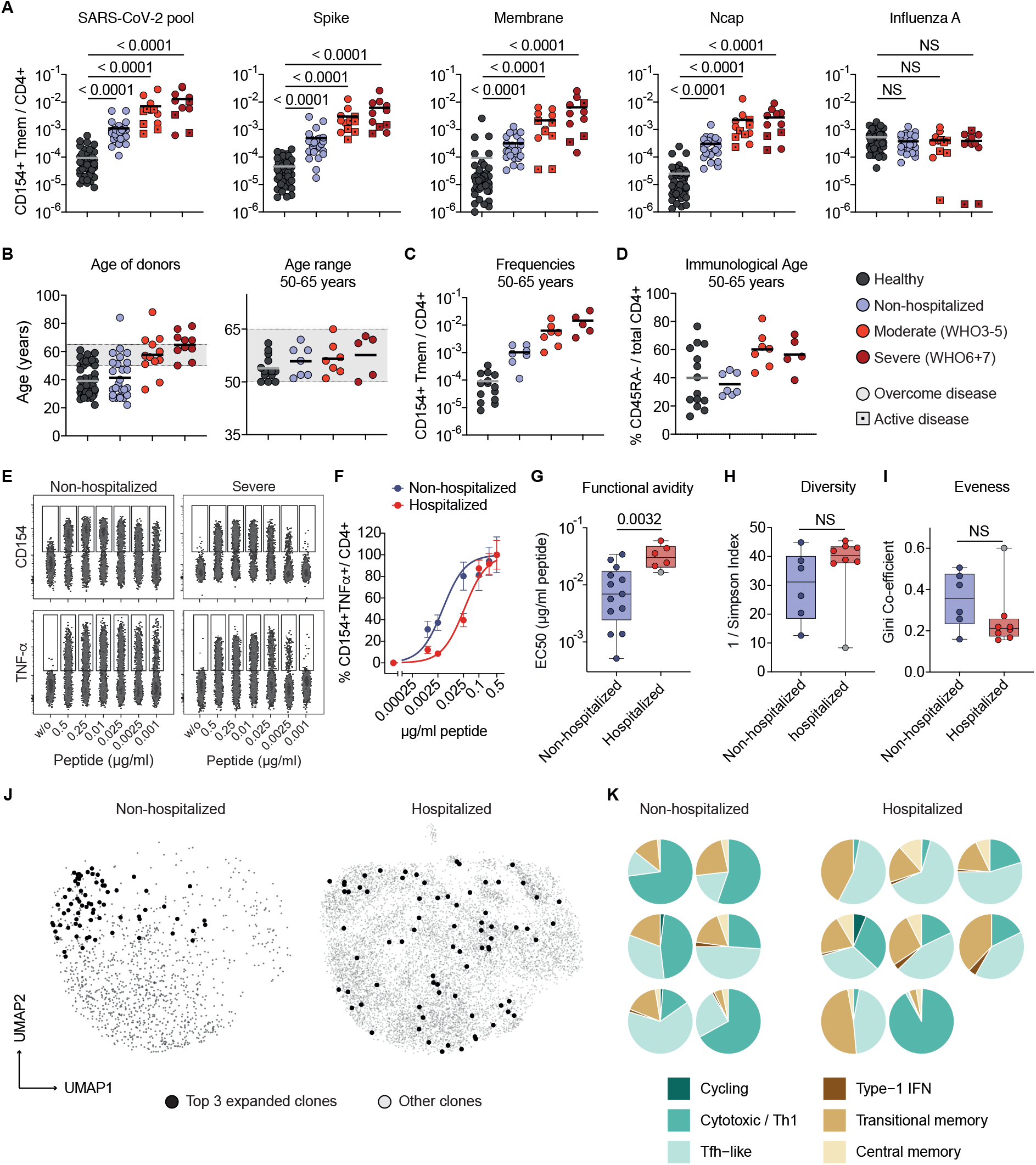
Unfocussed T cell response in severe COVID-19. (A) Frequencies of SARS-CoV-2-reactive Tmem. The highest COVID-19 severity level during disease was assessed based on WHO criteria, whereby WHO groups 3-5 (moderate) and 6-7 (severe) were combined to increase statistical power (see Table S1). Unexposed donors n=50, Non-hospitalized n=26 (WHO 1-2), mild-moderate n=12 (WHO 3 n=2, WHO 4 n=6, WHO 5 n=4), severe n=11 (WHO 6 n=5, WHO 7 n=6); patients with active disease at the time point of sampling are indicated with a square. (B) Age distribution within the different disease groups and controls and within the age-selected donors from 50-65 years. (C) Frequencies of SARS-CoV-2-pool-reactive Tmem in age-selected donors. (D) Immunological age of the age-selected donors, indicated as the proportion of memory cells within the total CD4+ population. (E-G) SARS-CoV-2 pool-reactive CD154+ Tmem were FACS purified, expanded and re-stimulated with decreasing antigen concentration in the presence of autologous antigen-presenting cells. (E) CD154 or TNF-a expression for the indicated concentration per peptide. (G) Dose-response curves of expanded T cell lines, restimulated with decreasing antigen concentrations. (F) EC50 values were calculated from dose-response curves. Non-hospitalized n=13, hospitalized n=6. (H, I) T cell receptor (TCR) sequence analysis from single cell data of the top 50 expanded clonotypes. (H) Simpson Index of TCR diversity. (I) Gini coefficient depicting the distribution of TCR sequences (0 is total equality, i.e. all clones have the same proportion, 1 total inequality, i.e. a population dominated by a single clone). Non-hospitalized n=6, hospitalized n=8. (J) Representative distribution of the top 3 expanded TCR clonotypes projected to the UMAP analysis for one exemplary non-hospitalized and one hospitalized COVID-19 patient. (K) Proportional distribution of the top 3 expanded clonotypes on the different Seurat clusters for each analyzed patient (non-hospitalized n=6; hospitalized n=8). Each symbol in (A-D, G-I) represents one donor, horizontal lines indicate (A-D) mean. (G-I) Box-and-whisker plots display quartiles and range. Statistical differences: (A) Kruskal-Wallis test with Dunn’s post hoc test, significant differences are indicated. (G-I) Two-tailed Mann-Whitney test.

To test whether the immunological age-related cross-reactive memory may impact on COVID-19 severity, we also compared TCR avidities and clonalities of SARS-CoV-2-reactive T cells from hospitalized *versus* non-hospitalized patients. Strikingly, SARS-CoV-2 reactive T cells from hospitalized patients displayed significantly lower functional avidity compared to non-hospitalized patients (Figure 6E-G). In line with this, SARS-CoV-2-specific T cells from hospitalized COVID-19 patients displayed a trend towards a more diverse TCR repertoire (Figure 6H) and reduced clonal expansions, as indicated by the lower Gini coefficient, as a measure of the eveness of a population (Figure 6I). However, this was not significant due to one outlier (grey dot in Figure 6G-I, see below). Thus despite strongly increased T cell frequencies in severe COVID-19 (Figure 6A), this increase did not result from an expansion of individual clones, but instead reflected a broad polyclonal response. We next analyzed the distribution of the most clonally expanded TCRs per patient within the different clusters of the single-cell RNA sequencing analysis. Interestingly, we observed a tendency that in mild disease the most expanded clones were mainly restricted to the cytotoxic cluster, whereas in more severe disease, they were scattered over several clusters (Figure 6J, K). One severe COVID-19 patient (grey dot in Figure 6G-I) did not fit into this scheme and also showed a high clonality strongly focused to the cytotoxic cluster (Figure 6K, lower right). Interestingly this patient suffered from a CMV reactivation, which may account for expansion of cross-reactive clones. Still the cells from this donor were of low avidity for SARS-CoV-2 antigens (Figure 6G, grey dot) confirming the robustness of the avidity data.

In summary our data suggest that severe COVID-19 disease is characterized by a strong but rather unfocused virus-specific CD4+ T cell response involving a broad polyclonal repertoire of rather low avidity T cells. Such unfocused, low avidity response may in fact result from preferential recruitment of a broad pre-existing memory repertoire preferentially present in the elderly.

## Discussion

Defining the parameters contributing to the high clinical variability of COVID-19 is essential to predict disease outcome and develop effective therapeutic and vaccination strategies. Here we provide two key observations, suggesting a negative impact of pre-existing T cell memory which may explain the age-bias of COVID-19 severity: First, we show that all COVID-19 patients generate strong pro-inflammatory T cells responses, that increased with disease severity. Unexpectedly, severe disease is associated with lower functional avidity and TCR clonality. Second, we identify SARS-CoV-2 “pre-existing” T cell memory as a common feature related to the immunological age of an individual, which recapitulates the low functional avidity found in severe COVID-19 and suggests a causative relation.

Our cytometric and single-cell RNA sequencing characterization of SARS-CoV-2 memory T cells confirmed previous results showing common characteristics of an anti-viral T cell response but did not identify clear-cut differences between severe and mild disease. Surprisingly, similar cell clusters were present in SARS-CoV-2-specific memory T cells from unexposed controls. Thus quantitative differences rather than unique functionality profiles develop in COVID-19. Indeed, all COVID-19 patients develop strong, pro-inflammatory Th1-like CD4+ T cell responses directed against the three main proteins spike, membrane and Ncap, as shown before for convalescent patients (Grifoni et al., 2020). Interestingly, despite the reported T cell lymphopenia in severe disease, SARS-CoV-2-specific T cell frequencies increased with disease severity (Anft et al., 2020; Peng et al., 2020). Compared to other common viruses SARS-CoV-2-specific T cells showed signs of recent activation, such as CD38, Ki-67 and PD1, as well as CD154 and high IL-21 production indicative of B cell helper function. Also the slightly reduced expression of cytokines like IFN-γ, TNF-a and IL-2 compared to other anti-viral responses may be related to the recent activation. Thus the T cell response phenotype *per se* does not explain disease severity but will require detailed longitudinal analysis in the future. The preferential formation of a highly focused clonal T cell population within the cytotoxic cluster in mild COVID-19 suggests their potential protective function, which may deserve further detailled analysis, including their peptide specificities.

We also characterized pre-existing memory as one factor for quantitative differences in the T cell response in mild versus severe disease. The observation that SARS-CoV-2 specific T cells were found in a subset of unexposed donors (Braun et al., 2020; Grifoni et al., 2020; Le Bert et al., 2020; Mateus et al., 2020; Meckiff et al., 2020; Sekine et al., 2020; Weiskopf et al., 2020), has initially fueled the hypothesis of protective pre-existing immunity, for example induced by related CCCoVs preferentially in young people (Braun et al., 2020; Mateus et al., 2020). Such heterologous immunity between related pathogens has been previously demonstrated mainly in infection models (Welsh et al., 2010) but may also modulate human immune responses (Bacher et al., 2019; Gras et al., 2010; Hayward et al., 2015; Koutsakos et al., 2019; Sridhar et al., 2013). So far, cross-reactivity of SARS-CoV-2 reactive T cells was poorly characterized and their functional impact remained unknown. In line with our results, a weak correlation between CCCoV and SARS-CoV-2 T cell responses in unexposed donors has been identified and cross-reactivity to CCCoV was observed in SARS-CoV-2-specific T cell lines directed against the most homologous part of spike proteins (Braun et al., 2020) or selected homologous peptides (Mateus et al., 2020). Our more detailled analysis of cross-reactivity in healthy and COVID-19 patients argues against a major role of CCCoVs: Pre-existing memory T cells were detected in all unexposed donors and their frequencies correlated with the immunological age but not with CCCoV-specific memory. Furthermore pre-existing memory cells displayed low functional avidity and were less focused on the dominant COVID-19 targets spike, membrane and Ncap protein (Figure 1D) (Grifoni et al., 2020; Le Bert et al., 2020). Most importantly, CCCoV cross-reactivity both within SARS-CoV-2-specific T cells from COVID-19 patients, as well as unexposed donors, was marginal despite the ubiquitous presence of a strong CCCoV-specific memory T cell response in all tested donors. Interestingly, also Mateus et al. found that SARS-CoV-2-specific, but not cross-reactive T cells against the homologous CCCoV peptides increased in COVID-19 patients (Mateus et al., 2020), supporting our finding. Thus CCCoV-specific T cell memory is common in the human population but seems to have minimal impact on SARS-CoV-2-specific immunity. It is important to mention that our demonstration of strong T cell memory against all four CCCoV strains in all tested donors may be an encouraging sign that protective cellular immunity against SARS-CoV-2 might also persist longterm, even if antibody responses are transient (Seow, 2020).

An even more important role of pre-existing memory in COVID-19 and for human immunity in general is emerging from our analysis. In contrast to previous reports we find memory T cells against SARS-CoV-2 in all tested unexposed donors. This probably reflects the high sensitivity and specificity of the ARTE assay. In fact, lack of magnetic pre-selection, prolonged stimulation times and the use of frozen PBMC may limit sensitivity and specificity (Bacher and Scheffold, 2013, 2015). However, this pre-existing memory does not represent classical heterologous immunity between related pathogens. Instead, pre-existing SARS-CoV-2 memory has features of an unbiased, stochastic cross-reactivity within a large TCR repertoire similar as observed against other neoantigens. This is supported by its ubiquitous presence and broad protein specificity (Figure 1D) as well as its strong positive correlation with total CD4+ memory (Figure 4D, E). Especially the low functional avidity argues against *in vivo* affinity selection (Figure 4G) (Bacher et al., 2016). Memory T cells against neo-antigens are commonly detected in humans (Bacher et al., 2013; Campion et al., 2014; Kwok et al., 2012; Su et al., 2013). This can be explained by the known TCR-intrinsic cross-reactivity against related and even unrelated but structurally similar peptides (Birnbaum et al., 2014; Sewell, 2012). Thus a highly diverse memory pool, which accumulates in humans over lifetime contains TCRs specific for neo-antigens similar to the naïve T cell pool.

The impact of pre-existing memory on T cell responses against neoantigens in humans is poorly understood. However, its correlation with the immunological age suggests increasing impact in the elderly (Lanzer et al., 2018; Lanzer et al., 2014; Woodland and Blackman, 2006) and it is tempting to speculate that this may contribute to the increased risk for severe COVID-19 in the aged population. Since memory T cells have a lower activation threshold, a large number of suboptimal low avidity memory cells may compete and prevent naïve T cell activation and high affinity selection (Lanzer et al., 2018). Indeed the size of the naïve T cell pool has been shown to correspond to vaccination success (Kwok et al., 2012; Schulz et al., 2015; Woodland and Blackman, 2006). Thus we hypothesize that pre-existing memory may contribute to the reduced avidity and higher diversity of TCRs in severe COVID-19. Such a polyclonal and low avidity T cell response may also be less susceptible to intrinsic negative control mechansims, which may explain the increased SARS-CoV-2-reactive T cell response. Pre-existing memory indeed represents a general mechanism of immune-modulation towards neo-antigens, especially in the elderly (Woodland and Blackman, 2006). Provided the great heterogeneity within the human population with regard to antigen exposure and MHC composition, we expect in fact highly variable and context-dependent effects of pre-existing memory from protective to harmful. Therefore the impact of pre-existing memory on neoantigen exposure, including sensitizing antigens, infections or vaccinations, as well as for autoantigens has to be carefully evaluated in future studies.

## Data Availability

Authors can confirm that all relevant data are included in the article and/or its supplementary information files

## Acknowledgments

We thank Chiara Romagnani and Ulf Klein for critical reading of the manuscript, the flow cytometry facility Cyto Kiel, especially Esther Schiminsky and the Competence Centre for Genomic Analysis (CCGA) Kiel, especially Janina Fuß, Sören Franzesnburg, Yewgenia Dolshanskaya, Catharina von der Lancken, Melanie Vollstedt and Melanie Schlapkohl for support with cell sorting and single cell sequencing; the Clinical Trial Unit 2 of the University Hospital of Cologne for help with recruiting study participants and technical assistance.

This research was supported by the German Research Foundation (DFG) under Germany’s Excellence Strategy - EXC 2167-390884018 Precision Medicine in Chronic Inflammation (to P.B., A.F., A.S.); RU5042 - miTarget to P.B., A.F.; DFG grant n. 4096610003 to E.R; FL and DE were supported by the E-Rare Joint Transnational research support (ERA-Net, LE3064/2-1).

## Author contributions

Conceptualization: P.B., A.S.; Investigation: P.B., E.R., G.R.M., C.S.; Formal analysis: P.B., E. R., D.E. P.K.; Resources: J.D., I.S., I.W., F.E., Y.K., M.J.G.T., C.C., F.T., P.R., R.M., K.P.W., F. L., J.R., M.K., O.A.C., P.K., A.F.; Funding acquisition: P.B., A.S.; All authors provided discussion, participated in revising the manuscript, and agreed to the final version.

---

**Figure S1.**
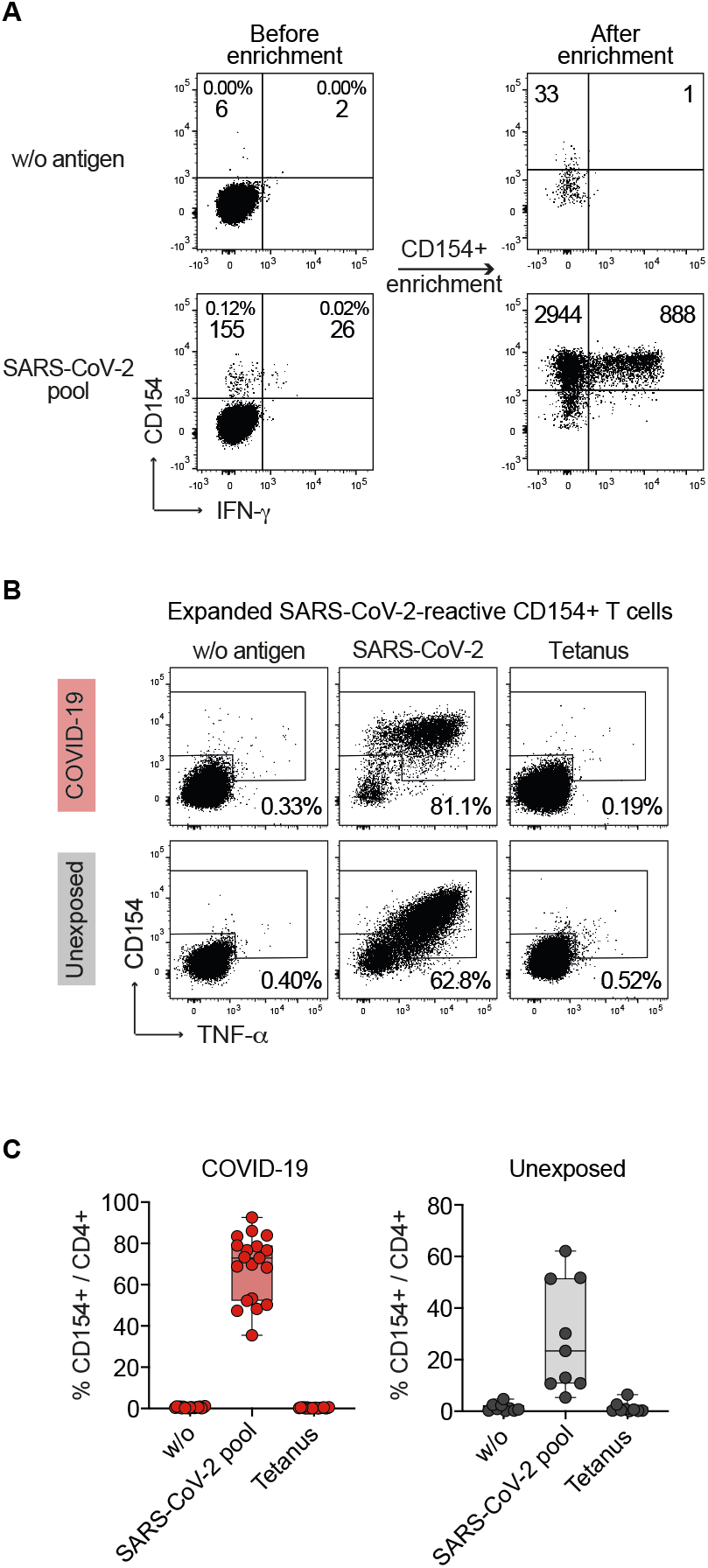
Detection of SARS-CoV-2 reactive CD4+ T cells by ARTE. (A) *Ex vivo* detection of SARS-CoV-2 pool-reactive CD4+ T cells by ARTE. Percentage within CD4+ T cells and absolute cell counts before and after magnetic CD154+ enrichment from 1×10e7 PBMCs are indicated. (B and C) Re-stimulation of FACS purified, expanded SARS-CoV-2 pool-reactive CD154+ T cells with the SARS-CoV-2 pool or Tetanus as control antigen. (B) Percentage of CD154+TNFα+ cells within CD4+ is indicated. (C) Statistical summary, each symbol represents one donor. Box-and-whisker plots display quartiles and range. Unexposed donors (n=9), COVID-19 patients (n=19).

**Figure S2.**
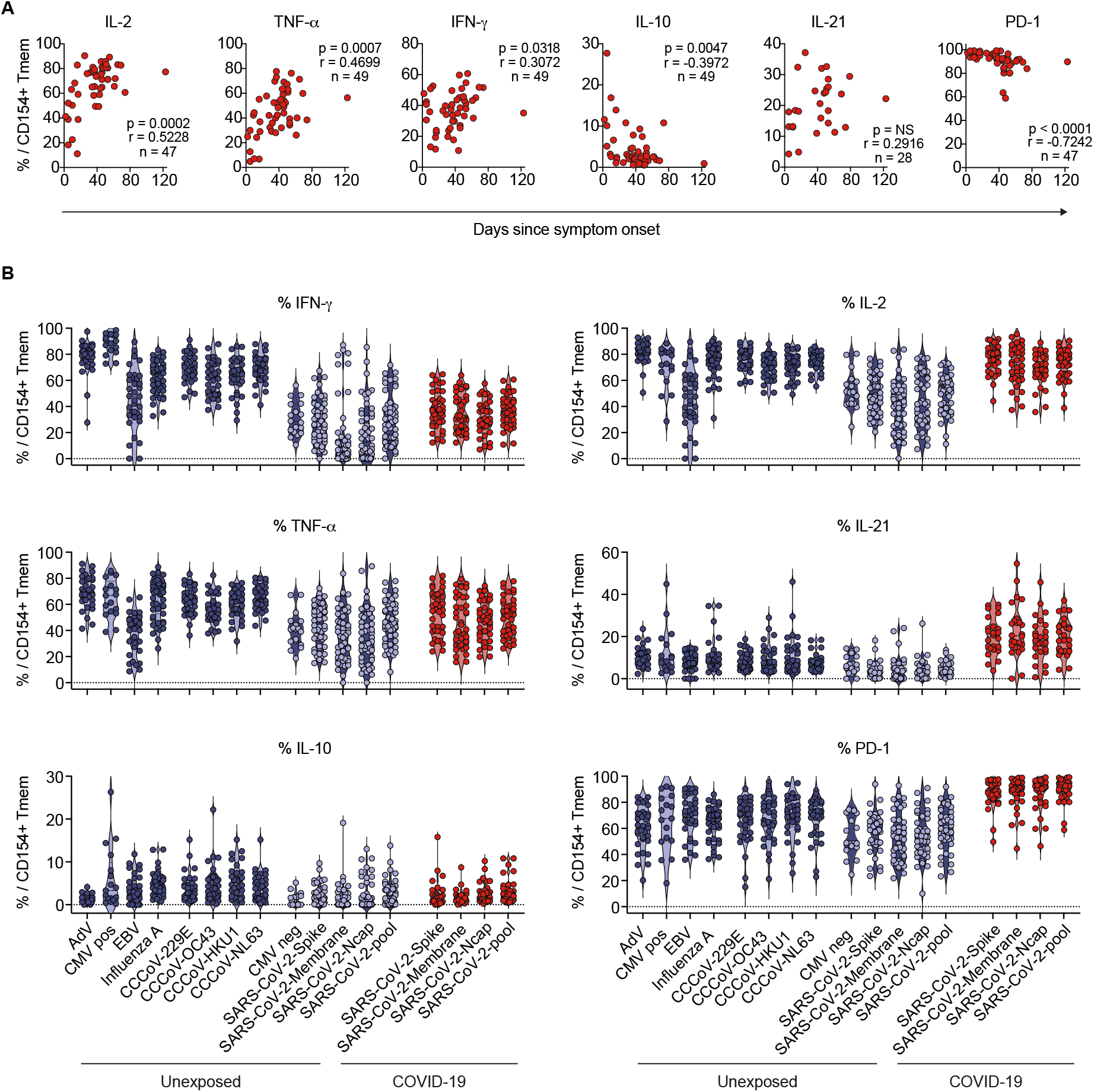
Pattern of SARS-CoV-2 reactive CD4+ T cells compared to other anti-viral responses. (A) Spearman correlation of cytokine and phenotypic marker expression of SARS-CoV-2 pool-reactive CD154+ Tmem and days since disease onset. (B) *Ex vivo* cytokine production and phenotype of SARS-CoV-2-reactive cells of reconvalescent COVID-19 patients in comparison to other anti-viral responses in SARS-CoV-2 unexposed donors (n=26-50). Each symbol in (A, B) represents one donor.

**Figure S3.**
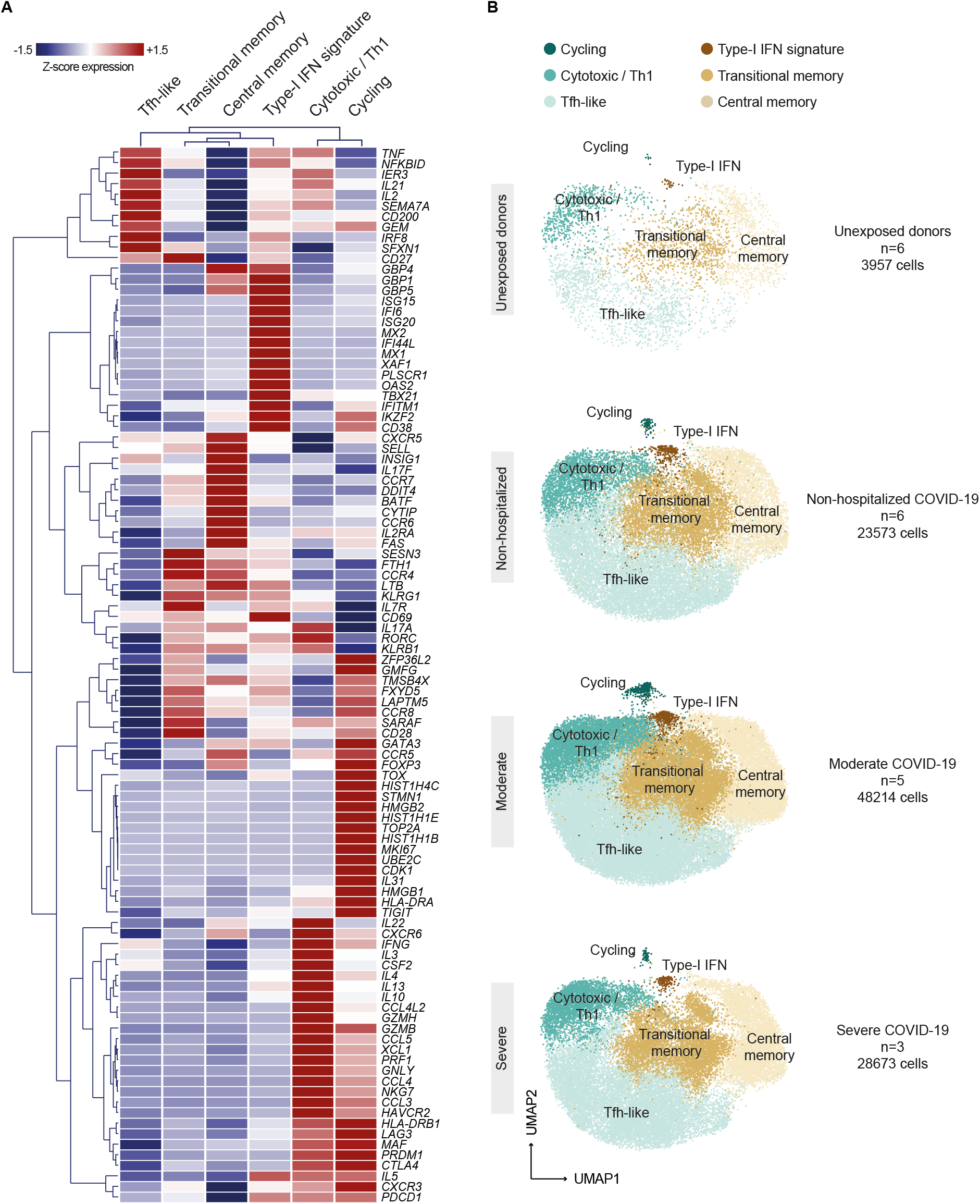
Gene expression of SARS-CoV-2 reactive CD4+ T cell clusters. Single cell transcriptomes of FACS purified *ex vivo* isolated CD154+ memory T cells following stimulation with pooled SARS-CoV-2 spike, membrane and Ncap proteins from unexposed donors (n=6) and COVID-19 patients (n=14). (A) Heatmap depicting Z-score normalized expression levels of the top 10 differential expressed marker genes of each cluster and other selected genes. (B) UMAP visualization of the subset composition of SARS-CoV-2 reactive CD4+ T cells colored by functional gene expression clusters for unexposed donors (n=6) and non-hospitalized (n=6), moderate (WHO 4-5; n=5) and severe (WHO 6-7; n=3) COVID-19 patients.

**Figure S4.**
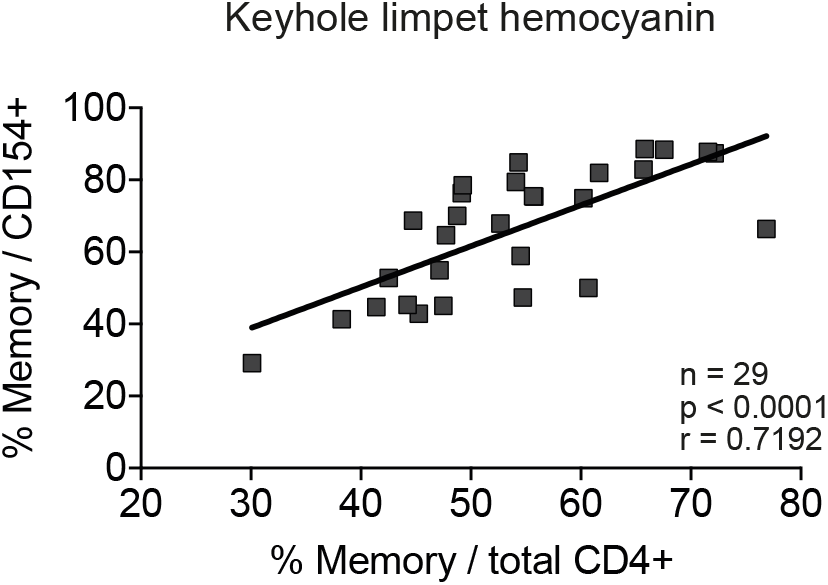
Proportion of neoantigen-specific memory T cells correlates with the immunological age. Pearson correlation between the proportion of memory cells within the antigen-specific T cells (y-axis) and the proportion of memory cells within the total CD4+ population is shown for the neoantigen keyhole limpet hemocyanin (KLH).

**Figure S5.**
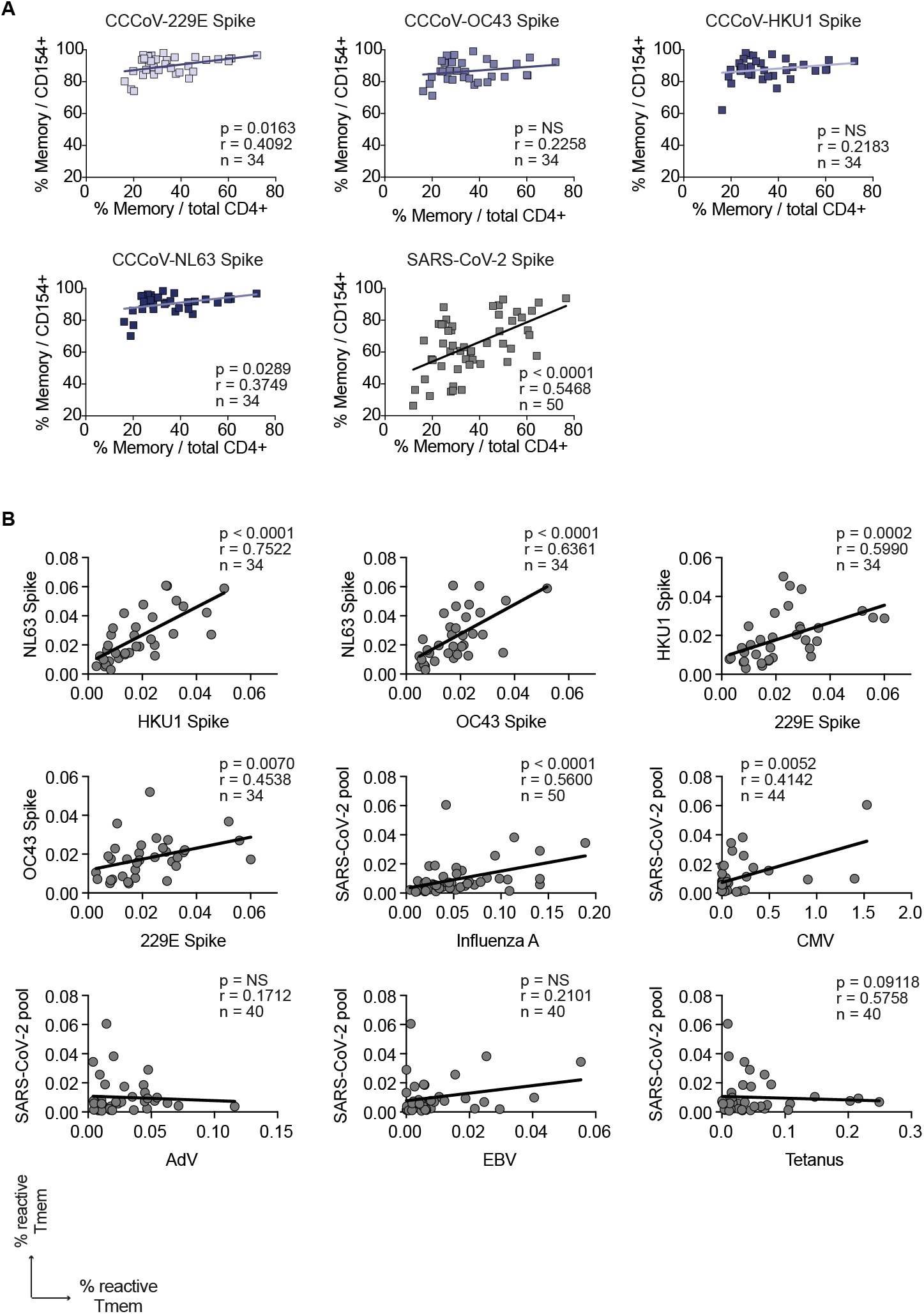
Correlations of SARS-CoV-2-reactive T cells of unexposed donors with the response against other common viruses. (A) Pearson correlation between the proportion of memory cells within the CCCoV spike-specific T cells (y-axis) and the proportion of memory cells within the total CD4+ population (x-axis, immunological age) in SARS-CoV2-unexposed donors. (B) Spearman correlation between CD154+ Tmem frequencies reactive against different cCcoVs or SARS-CoV-2 and Influenza A (H1N1), Cytomegalovirus (CMV), Epstein–Barr Virus (EBV), Adenovirus (AdV) or tetanus in unexposed donors. Each symbol in (A, B) represents one donor.

**Figure S6.**
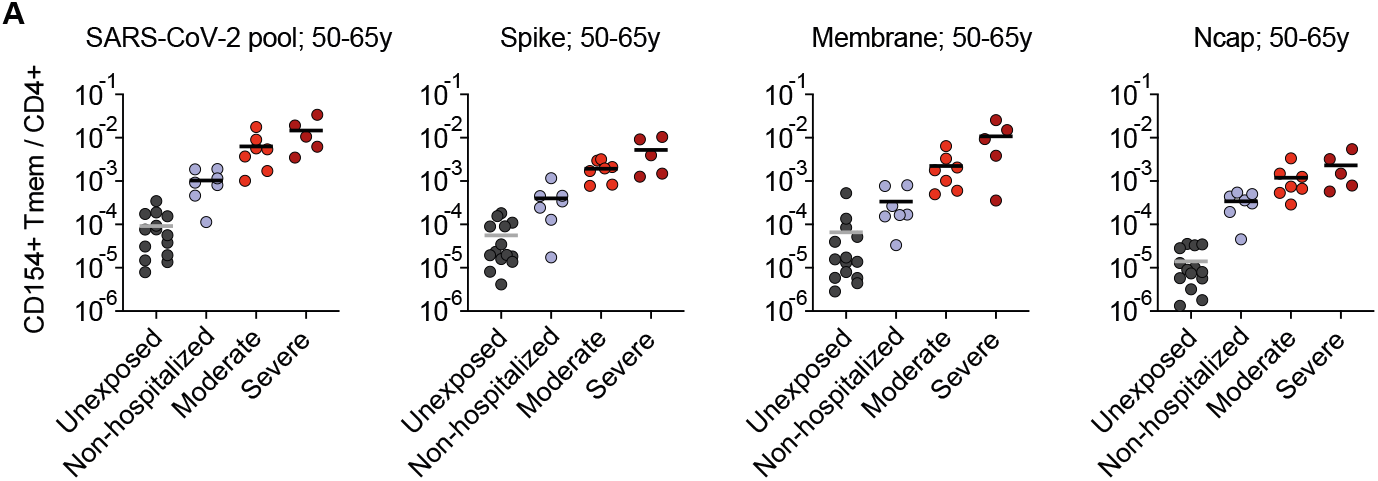
SARS-CoV-2-reactive T cell in age-selected donors. (A) Frequencies of Tmem reactive aganst the indicated SARS-CoV-2 proteins in donors with an age range of 50-65. The highest COVID-19 severity level during disease was assessed based on WHO criteria, whereby WHO groups 3-5 (moderate) and 6-7 (severe) were combined to increase statistical power (see table S1). Unexposed donors n=14, Non-hospitalized n=7 (WHO 1-2), moderate n=7 (WHO 3 n=1, WHO 4 n=3, WHO 5 n=3), severe n=5 (WHO 6 n=2, WHO 7 n=3,). Each symbol in represents one donor, horizontal lines indicate mean.

## Materials & Methods

### CONTACT FOR REAGENT AND RESOURCE SHARING

Further information and requests for reagents may be directed to the corresponding author Petra Bacher.

## EXPERIMENTAL MODEL AND SUBJECT DETAILS

### COVID-19 patients and unexposed donors

This study was approved by the Institutional Review board of the UKSH Kiel (Identifier D 474/20), the University Hospital Frankfurt (Identifier 11/17) and patients were enrolled in the protocol Coronavirus Disease 19 – BioMaSOTA - Genetic factors and longitudinal monitoring of the immune response in COVID-19 (Identifier of the University of Cologne Ethics Committee 20-1295) and Improving Diagnosis of Severe Infections of Immunocompromised Patients (Identifier of the University of Cologne Ethics Committee 08-160) and signed informed consents.

Peripheral EDTA blood samples were collected between April and July 2020 from 49 COVID-19 patients and from 50 in-house volunteers as unexposed controls (Table S1). 44 of 49 COVID-19 patients were tested positive and for SARS-CoV-2 RNA. We included 5 mild cases of COVID-19 without positive SARS-CoV2 RNA test, but with positive detection of antibodies using a certified antibody test (Elecsys Anti-SARS-CoV-2, Roche Diagnostics GmbH, Mannheim, Germany) who had clinical symptoms suggestive of COVID-19 and a traceable contact person found positive.

All, except three active COVID-19 patients who had a positive SARS-CoV-2 RNA test, were tested positive for SARS-CoV-2 antibodies (Elecsys Anti-SARS-CoV-2, Roche Diagnostics GmbH and/ or Anti-SARS-CoV-2 ELISA, Euroimmun, Lübeck, Germany). All healthy controls were tested negative for SARS-CoV-2 antibodies (Elecsys Anti-SARS-CoV-2, Roche Diagnostics GmbH). The highest COVID-19 severity was assessed based on WHO ordinal scale (https://www.who.int/publications/i/item/covid-19-therapeutic-trial-synopsis).

## METHOD DETAILS

### Antigens

Pools of lyophilized 15-mer peptides with 11–amino acid overlap, covering the complete protein sequence were purchased from Miltenyi Biotec (Bergisch Gladbach, Germany): SARS-CoV-2 Membrane, Ncap or JPT (Berlin, Germany): SARS-CoV-2 Spike N-term, Spike C-term, AP3A, ORF9B, ORF10, NS6, NS7a, NS7b, NS8, VEMP, Y14.

Peptide pools of control antigens Influenza A H1N1 (HA, MP1, MP2, NP and NA), CMV (pp65, IE-1), EBV (EBNA1, BZLF1, LMP2A, LMP1), AdV (Hexon) were purchased from Miltenyi Biotec and CCCoV Spike proteins (229E, OC43, HKU1, NL63) from JPT. Pools were resuspended according to manufacturer’s instructions and cells were stimulated at a concentration of 0.5 μg/peptide/ml. Tetanus-toxoid was purchased from Statens Serum Institute and used at a concentration of 10μg/ml.

### Antigen-reactive T cell enrichment (ARTE)

Peripheral blood mononuclear cells were freshly isolated from 20-50ml EDTA blood on the day of blood donation by density gradient centrifugation (Biocoll; Biochrom, Berlin, Germany). Antigen-reactive T cell enrichment (ARTE) was performed as previously described (Bacher et al., 2019; Bacher et al., 2016). In brief, 0.5-2×10e7 PBMCs were plated in RPMI-1640 medium (GIBCO), supplemented with 5% (v/v) human AB-serum (Sigma Aldrich, Schnelldorf, Germany) at a cell density of 1×10e7 PBMCs / 2 cm^2^ in cell culture plates and stimulated for 7 hr in presence of 1 μg/ml CD40 and 1 μg/ml CD28 pure antibody (both Miltenyi Biotec, Bergisch Gladbach, Germany). 1 μg/ml Brefeldin A (Sigma Aldrich) was added for the last 2 hr.

Cells were labeled with CD154-Biotin followed by anti-Biotin (CD154 MicroBead Kit, Miltenyi Biotec) and magnetically enriched by two sequential MS columns (Miltenyi Biotec). Surface staining was performed on the first column, followed by fixation and intracellular staining on the second column. Frequencies of antigen-specific T cells were determined based on the cell count of CD154+ T cells after enrichment, normalized to the total number of CD4+ T cells applied on the column. For each stimulation, CD154+ background cells enriched from the non-stimulated control were subtracted.

### Flow cytometry

Cells were stained in different combinations of fluorochrome-conjugated antibodies (see Key Resources Table). Viobility 405/520 Fixable Dye (Miltenyi Biotec) was used to exclude dead cells. For intracellular staining cells were fixed and permeabilized with the Inside stain Kit (Miltenyi Biotec). Data were acquired on a or LSR Fortessa (BD Bioscience, San Jose, CA, USA). Screening of expanded T cell lines on 384-well plates was performed on a MACSQuantX Analyzer (Miltenyi Biotec). FlowJo (Treestar, Ashland, OR, USA) software was used for analysis.

### *In vitro* expansion and re-stimulation of antigen-reactive T cell lines

For expansion of antigen-specific T cell lines, PBMCs were stimulated for 6 hr, CD154+ cells were isolated by MACS and further purified by FACS sorting on a FACS Aria Fusion (BD Bioscience, San Jose, CA, USA) based on dual expression of CD154 and CD69. Purified CD154+ T cells were expanded in presence of 1:100 autologous antigen-loaded irradiated feeder cells in TexMACS medium (Miltenyi Biotec), supplemented with 5% (v/v) human AB-serum (GemCell), 200 U/ml IL-2 (Proleukin; Novartis, Nürnberg, Germany), and 100 IU/ml penicillin, 100 μg/ml streptomycin, 0.25 μg/ml amphotericin B (Antibiotic Antimycotic Solution, Sigma Aldrich) at a density of 2.5×106 cells/cm^2^. During expansion for 2-3 weeks, medium was replenished and cells were split as needed.

For re-stimulation, fastDCs were generated from autologous CD14+ MACS isolated monocytes (CD14 MicroBeads; Miltenyi Biotec) by cultivation in X-Vivo™15 medium (BioWhittaker/Lonza), supplemented with 1000 IU/ml GM-CSF and 400 IU/ml IL-4 (both Miltenyi Biotec). Before re-stimulation expanded T cells were rested in RPMI-1640 + 5 % human AB-serum without IL-2 for 2 days. 0.5-1×10e5 expanded T cells were plated with fastDCs in a ratio 1:1 of in 384-well flat bottom plates and re-stimulated for 6 h, with 1 μg/ml Brefeldin A (Sigma Aldrich) added for the last 4 hr.

### Cell isolation and single-cell RNA-seq assay (10x Genomics)

For single cell transcriptomics, CD154+ cells were isolated by MACS and further purified by FACS sorting on a MACSQuant Tyto (Miltenyi Biotec) based on dual expression of CD154 and CD69. Sorted CD154+ T cells were removed from the sorting chamber into pre-coated low-bind collection tubes, 1ml RPMI1640 medium supplemented with 5% AB Serum was added, and cells were centrifuged for 5 min at 400 × g, 4°C. The supernatant was carefully removed leaving 10-30μl to reach a maximum concentration of 1000 cells /μl.

Single-cell suspensions were loaded on a Chromium Chip G (10x Genomics) according to the manufacturer’s instructions for processing with the Chromium Next GEM Single Cell 5’ Library and Gel Bead Kit v1.1. Depending on the number of cells available for each patient, a maximum of 30,000 cells were loaded for each reaction. TCR single-cell libraries were subsequently prepared from the same cells with the Chromium Single Cell V(D)J Enrichment Kit, Human T Cell. Libraries were sequenced on Illumina NovaSeq 6000 machine with 2×100 bp for gene expression, aiming for 50,000 reads per cell and 2×150 bp and 5000 reads per cell for TCR libraries.

### Single cell T cell receptor (TCR) sequence analysis

Single-cell T-cell receptor repertoire clonotype tables were generated using the VDJ command of the Cellranger software, version 3.1.0. from 10xGenomics and using the reference GRCh38 version 2.0.0. Clonotype tables were filtered in order to include only cells which passed quality filtering in the gene expression analysis. In addition, clonotypes were stringently filtered for possible doublets by removing clonotypes (i) found in 1 cell only and containing more than 1 TCR alpha and 1 TCR beta (ii) containing more than 1 TCR alpha and no TCR beta sequence (iii) containing more than 1 TCR beta and no TCR alpha sequence (iv) containing more than 2 TCR alpha or more than 2 TCR beta sequences.

Alpha diversity measures were calculated for each patient either for the whole repertoire or divided based on Seurat clusters. R packages “vegan” and “tcR” were used to calculate the Inverse Simpson diversity index and the Gini inequality index, respectively. For these analyses samples were normalized by selection of the most abundant 50 clonotypes in order to remove the impact of different sample sizes (number of cells per sample) and to analyze only the distribution of the most expanded clonotypes.

Analysis of the most expanded clonotypes was conducted by selecting the 3 most expanded clonotypes per sample. To evaluate potentially existing preferential cumulation of most expanded clonotypes in certain functional clusters, the proportion of cells carrying these clonotypes falling in each distinct Seurat cluster was calculated.

### Single-cell transcriptome analysis

The preprocessing of the scRNA-data was performed with the 10x Genomics’ Cell Ranger software v3.1.0 using the reference GRCh38 v3.0.0 for the mappings. The resulting filtered feature-barcode matrix files were analyzed with the R package Seurat v.3.2.0 (Butler et al., 2018). Thereby, all genes with a detected expression in less than 0.1% of the non-empty cells were excluded. Moreover, TCR genes were not considered for further analyses to avoid functional clustering of cells based on TCR information. To minimize the number of doublets, empty cells, and cells with a transcriptome in low quality, only cells harboring between 840 (minimum median among samples) and 3000 RNA features and less than 5% mitochondrial RNA were selected for further processing. Afterwards, data were log-normalized and scaled based on all genes. After performing a PCA dimensionality reduction (20 dimensions) with the RunPCA function, the expression values were corrected for effects caused by different sample preparation time points in time using the R package Harmony v1.0 (Korsunsky et al., 2019). In the final steps, the Uniform Manifold Approximation and Projection (UMAP) dimensional reduction was performed with the RunUMAP function using 20 dimensions, a shared nearest neighbor graph was created with the FindNeighbors method, and the clusters identification was performed with a resolution of 0.2 using the FindClusters function. Positive cluster marker genes were determined using FindMarkers with the MAST method (Finak et al., 2015). Thereby, only genes with detected expression in at least 25% of the cells in the respective cluster were considered.

## QUANTIFICATION AND STATISTICAL ANALYSIS

Statistical parameters including the exact value of n, the definition of center, dispersion and precision measure, and statistical significance are reported in the Figures and the Figure Legends. Statistical tests were performed with GraphPad PRISM software 8.4 (GraphPad Software, La Jolla, CA, USA). Statistical tests were selected based on appropriate assumptions with respect to data distribution and variance characteristics, p values < 0.05 were considered statistically significant.

## DATA AND SOFTWARE AVAILABILITY

### Software

Flow-cytometry data were analyzed using FlowJo (Treestar, Ashland, OR, USA) software. Graphics and statistics were created with GraphPad PRISM software version 8.4.3. (GraphPad Software, La Jolla, CA, USA). Heatmaps were generated using Genesis software (Sturn et al., 2002), version 1.7.7.

**Table S1.**
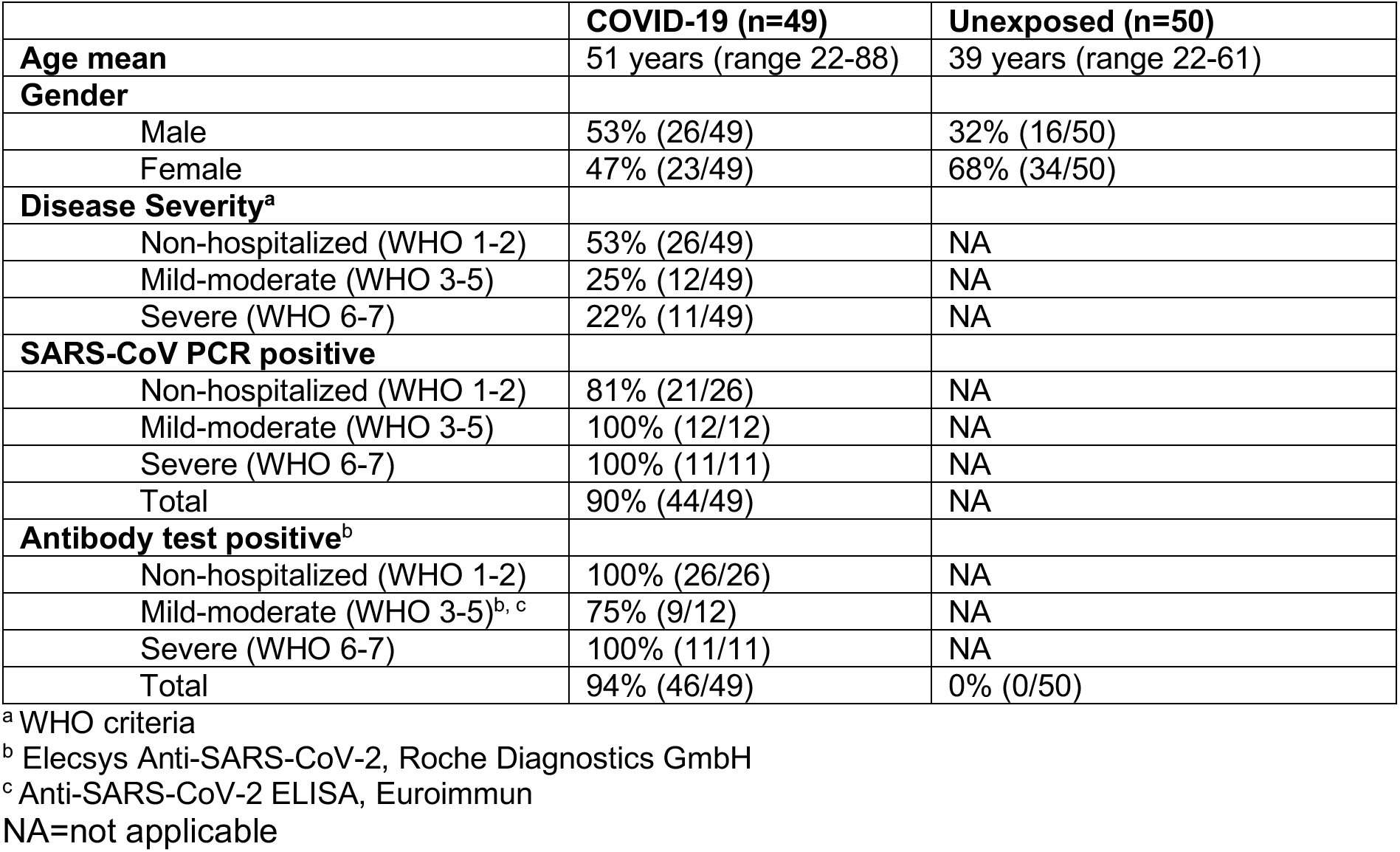
Cohort characteristics.

|  | **COVID-19 (n=49)** | **Unexposed (n=50)** |
| --- | --- | --- |
| **Age mean** | 51 years (range 22-88) | 39 years (range 22-61) |
| **Gender** |  |  |
| Male | 53% (26/49) | 32% (16/50) |
| Female | 47% (23/49) | 68% (34/50) |
| **Disease Severity<sup>a</sup>** |  |  |
| Non-hospitalized (WHO 1-2) | 53% (26/49) | NA |
| Mild-moderate (WHO 3-5) | 25% (12/49) | NA |
| Severe (WHO 6-7) | 22% (11/49) | NA |
| **SARS-CoV PCR positive** |  |  |
| Non-hospitalized (WHO 1-2) | 81% (21/26) | NA |
| Mild-moderate (WHO 3-5) | 100% (12/12) | NA |
| Severe (WHO 6-7) | 100% (11/11) | NA |
| Total | 90% (44/49) | NA |
| **Antibody test positive<sup>b</sup>** |  |  |
| Non-hospitalized (WHO 1-2) | 100% (26/26) | NA |
| Mild-moderate (WHO 3-5)<sup>b, c</sup> | 75% (9/12) | NA |
| Severe (WHO 6-7) | 100% (11/11) | NA |
| Total | 94% (46/49) | 0% (0/50) |

<sup>a</sup> WHO criteria<sup>b</sup> Elecsys Anti-SARS-CoV-2, Roche Diagnostics GmbH<sup>c</sup> Anti-SARS-CoV-2 ELISA, Euroimmun
NA=not applicable

